# Pulmonary radiological change of COVID-19 patients with ^99m^Tc-MDP treatment

**DOI:** 10.1101/2020.04.07.20054767

**Authors:** Xiaolin Yuan, Wanrong Yi, Baoyi Liu, Simiao Tian, Fang Cao, Ruoyu Wang, Baiwen Qi, Faqiang Lu, Meiyun Fang, Fuyang Pei, Ming Chen, Lichuan Zhang, Yong Zhang, Xiuzhi Zhang, Zhenyu Pan, Dewei Zhao, Aixi Yu

**Author notes:** **Corresponding author:** Dr. Dewei Zhao and Dr. Aixi Yu, Address correspondence to Dr. Dewei Zhao at Department of Orthopaedics, Affiliated Zhongshan Hospital of Dalian University, NO. 6 Jiefang Street Zhongshan District, Dalian, Liaoning Province, China, 116001., Address correspondence to Dr. Aixi Yu at Department of Orthopedics Trauma and Microsurgery, Zhongnan Hospital of Wuhan University, Wuhan, Hubei, 430071, China. contributed equally (Xiaolin Yuan, Wanrong Yi, Baoyi Liu).

## Abstract

**Background:** As increasing cases of COVID-19 around world, urgent need for effective COVID-19-specific therapeutic drugs is necessary; therefore, we conducted a pilot randomized-controlled study to evaluate the efficacy of ^99m^Tc-MDP for COVID-19 therapeutic treatment.

**Methods:** A total of 21 mild patients with COVID-19 were enrolled in this pilot RCT from February 2020 through March 2020, and then were assigned, in a 1:1 ratio, into control (11 patients) and ^99m^Tc-MDP group (10 patients). Patients in the control group received routine treatment and patients assigned to the ^99m^Tc-MDP group received a combination of routine treatment and an administration of ^99m^Tc-MDP injection of 5ml/day. Both of the patients in the control and ^99m^Tc-MDP groups were treated for 7 days with the primary end point of CT-based radiological pulmonary changes during 7-day follow-up.

**Findings:** From baseline to the day 7, 8 (80%) of 10 mild patients in the ^99m^Tc-MDP group had a significant radiological improvement in lung and a decline in inflammatory infiltration, whereas only 1 (9.1%) of 11 patients in the control group had a radiological improvement in lung. None of the patients in the ^99m^Tc-MDP group had disease progression from mild to severe, as well as an inflammatory cytokine storm, and 2 mild patients (18.2%) in the control group developed severe. During days 7 through 14, the number of patients with radiological improvement in the ^99m^Tc-MDP group remained consistent, and only 1 additional case (22%) in the control group were reported.

**Conclusion:** In this randomized pilot study, ^99m^Tc-MDP had an effective inhibitory effect on the inflammatory disease progression for the therapy of COVID-19, and it can accelerate the absorption of pulmonary inflammation in a short period of time during the process of treatment.

---

The recent outbreak of the novel coronavirus (2019-nCoV) is currently beginning spread to many countries around the world, and is becoming a pandemic according to World Health Organization (WHO).^1^ The increasing cases and fatality highlight the urgent need for effective COVID-19-specific therapeutic drugs, which would prevent the progression of the disease, especially for decreasing the deterioration rate of respiratory and pulmonary infection. In the process of COVID-19, the cytokine storm is trigged by excessive inflammatory response and macrophage activation, which lead to multiple organ damages and even fatality; therefore, suppressing inflammatory response and targeting macrophage are both crucial for COVID-19 treatment. ^99m^Tc-methylene diphosphonate (^99m^Tc-MDP) is a first-in-class, highly active inhibitor of inflammation response and macrophage infiltration, and it is common drug therapy for use in autoimmune diseases such as rheumatoid arthritis with no significant adverse effects in clinical use.^2^. In this pilot randomized controlled trial (RCT) study, ^99m^Tc-MDP were administered in combination, with or without standard treatment for 7 days in patients with COVID-19. We report preliminary results from this exploratory cohort, with main focus of computed tomography (CT)-based radiological pulmonary changes at 7 days.

A total of 21 mild patients with COVID-19 were enrolled in this pilot RCT from February 2020 through March 2020 (Trial Registration: ChiCTR2000029431). Patients were considered eligible in this study on the basis of the WHO interim guidance^3^ and they were laboratory-confirmed positive on by the means of real-time reverse-transcriptase–polymerase-chain-reaction (RT-PCR) assay of nasal and pharyngeal swab specimens. Randomization was performed and ensured with the use of Web-based randomization system. Patients were assigned, in a 1:1 ratio, into control (11 patients) and ^99m^Tc-MDP group (10 patients). Patients in the control group received routine treatment according to *Diagnostic of COVID-19* criteria issued by National Health Commission of China. Patients assigned to the ^99m^Tc-MDP group received a combination of routine treatment and an administration of ^99m^Tc-MDP injection of 5ml/day. Both of the patients in the control and ^99m^Tc-MDP groups were treated for 7 days. The primary end point was the CT-based radiological pulmonary changes from baseline to 7 days. The secondary end points were disease progression from mild to severe on the day 7, as well as radiological pulmonary changes during 7 days to 14 days after treatment.

Demographic characteristics were similar in the two groups except that patients in the control group were older than those in the ^99m^Tc-MDP group (median age, 62 years vs. 57 years), there were fewer women in the control group than in the ^99m^Tc-MDP group (55 % vs. 60%) (Table 1). On the day 7, 8 (80%) of 10 mild patients in the ^99m^Tc-MDP group had a significant radiological improvement in lung and a decline in inflammatory infiltration, whereas only 1 (9.1%) of 11 patients in the control group had a radiological improvement in lung (Figure 1 and Figure S1 in the Supplementary Appendix). None of the patients in the ^99m^Tc-MDP group had disease progression from mild to severe, as well as an inflammatory cytokine storm, and 2 mild patients (18.2%) in the control group developed severe. During days 7 through 14, the number of patients with radiological improvement in the ^99m^Tc-MDP group remained consistent, and only 1 additional case (22%) in the control group were reported (Table 1).

**Table 1.**
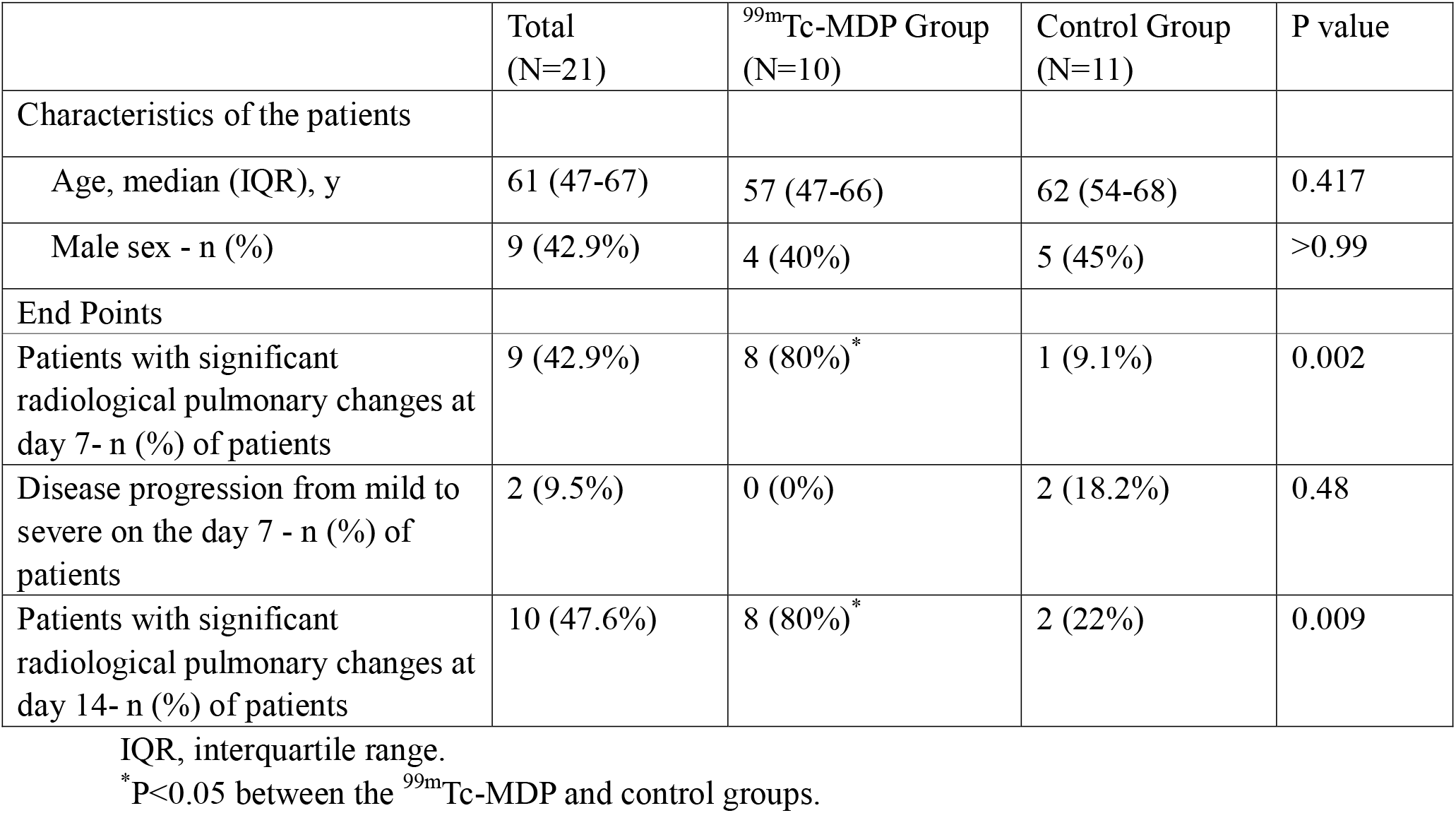
Characteristics of the patients and End Points

**Figure 1:**
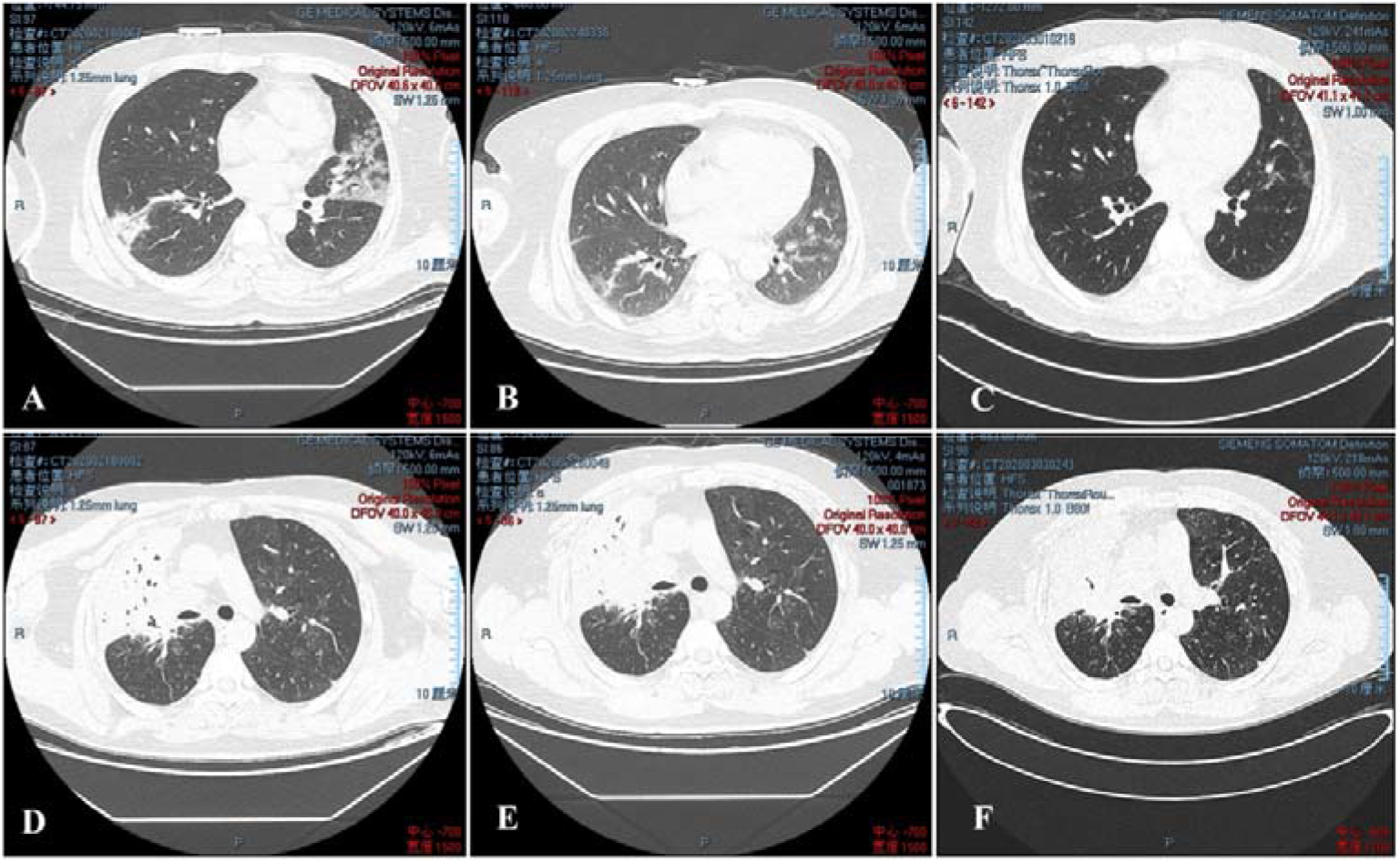
Computed Tomography (CT)-based radiographs of the patients’ lung. On the top, shown are pulmonary radiographs for one patient in the ^99m^Tc-MDP group at admission (A), at day 7 (B) and at day 14 (C); on the bottom, shown are pulmonary radiographs for one patient in control group at admission (D), at day 7 (E) and at day 14 (F).

In conclusion, in this randomized pilot study, the clinical drug, ^99m^Tc-MDP had an effective inhibitory effect on the inflammatory disease progression from mild to severe for the therapy of COVID-19, and it can accelerate the absorption of pulmonary inflammation in a short period of time during the process of treatment. Besides, there were no serious adverse events including body immunity during the clinical use of ^99m^Tc-MDP. Additional potential advantage for application of ^99m^Tc-MDP is to avoid excessive use of glucocorticoids in the treatment of severe COVID-19 patients, in order to prevent further incidence of some glucocorticoids caused complications such as osteonecrosis of femoral head.^4^ Limited by the small sample size, larger and longer trials are warranted to determine the efficacy of ^99m^Tc-MDP in the treatment of COVID-19.

## Data Availability

Data can be obtained if request.

## Notes

### Competing Interest Statement

The authors have declared no competing interest.

### Clinical Trial

ChiCTR2000029431

